# Automated Digital Biomarker Discovery Pipeline for Cardiovascular Diseases

**DOI:** 10.1101/2025.01.03.25319955

**Authors:** Gouthamaan Manimaran, Sadasivan Puthusserypady, Helena Dominguez, Jakob E. Bardram

## Abstract

Cardiovascular Diseases (CVDs) are the leading cause of mortality worldwide, necessitating early and accurate diagnosis to prevent severe outcomes such as Heart Failure (HF). Despite the widespread use of Electrocardiogram (ECG) for cardiac monitoring, traditional methods often miss subtle preclinical changes. In this paper, we present an automated digital biomarker discovery pipeline that leverages explainable artificial intelligence (XAI) to enhance the interpretability and clinical applicability of ECG-based biomarkers for CVDs. Using an interpretable feature extractor combined with unsupervised clustering and Particle Swarm Optimisation (PSO), our method identifies both known and novel ECG features associated with high CVD risk. These include established markers like RR Interval Sample Entropy and the discovery of novel biomarkers such as T-Wave Multiscale Entropy, which we found to be significantly associated with CVD risk. Our pipeline enhances early detection by bridging Artificial Intelligence (AI) methods with clinical relevance, providing interpretable insights that align with physiological principles. This transparency promotes clinician trust and supports the integration of AI into routine medical practice. Our results demonstrate that this approach can significantly improve the prediction and understanding of heart diseases, thus offering a powerful tool for reducing the global burden of CVDs.

## 1 Introduction

Cardiovascular Diseases (CVDs) are the leading cause of illness and death worldwide, significantly impacting global health. In 2021, CVDs accounted for approximately 21 million deaths, marking a 60% increase from approximately 12 million in 1990 [1]. Among these conditions, Heart Failure (HF) is particularly concerning because it often progresses without noticeable symptoms until it reaches an advanced stage [2]. This silent progression underscores the critical need for robust and precise methods to detect heart problems early, enabling timely interventions that can significantly improve patient outcomes.

Electrocardiogram (ECG) is the widely used, non-invasive tool for monitoring the heart’s electrical activity. Despite its widespread use, traditional ECG analysis often misses the subtle and complex changes in the heart that occur before clear symptoms of HF and other CVDs appear [3]. This gap underscores the necessity for more advanced methods to identify high-risk patients before significant heart damage occurs.

Recently, Artificial Intelligence (AI) has become a powerful tool in medical diagnostics, helping to analyze large and complex datasets. Deep learning models, in particular, have shown an impressive ability to interpret ECG data, sometimes even outperforming human experts [4]. There have been impressive work on CVD risk stratification using AI, where many works [5–7] try to predict the biological age of the patient using an end-to-end deep learning network and map the error in the predicted age to CVD risk. There have also been works [8, 9] that directly map 3, 5-year CVD risk to the ECG signal. However, a major challenge is that many such AI models act like “black boxes”, providing little insight into how they make their predictions. This lack of transparency makes it hard for doctors to trust these models and poses challenges to their approval and ethical use in healthcare, leading to the need for explainable AI (XAI) [10].

To address the lack of transparency and explainability in AI-based diagnosis in cardiology, this paper presents a new method to discover interpretable digital biomarkers from ECG data. We use the term ‘Digital Biomarker’ to denote human-understandable features, which are highly important in the automated AI models. By extracting such digital biomarkers from the ECG signal, these biomarkers can be presented to the medical doctors and help explain what features in the underlying signal, the AI algorithm uses for its prediction and diagnosis. Such biomarkers also make it possible for medical doctors and researchers to talk about, and share, diagnostics reasoning with each other and thus may become standards in CVD diagnosis.

Our approach uses a handcrafted feature extraction pipeline, combined with clustering algorithms, an objective function, and a search space optimization technique to find meaningful patterns that distinguish at-risk patients from healthy individuals. Unlike traditional black-box models, this method emphasizes transparency by allowing doctors to see and understand the significance of the identified biomarkers. By applying the proposed method, we demonstrate its ability to discover biomarkers that are strongly linked to HF and other CVDs. This includes identifying known biomarkers like ‘RR Interval Sample Entropy’ [11] and uncovering novel ones such as ‘T-Wave Multiscale Entropy’. In this paper, we present an overview of digital biomarkers for CVD and HF that have been discovered by using our discovery pipeline (see Table 1).

**Table 1:**
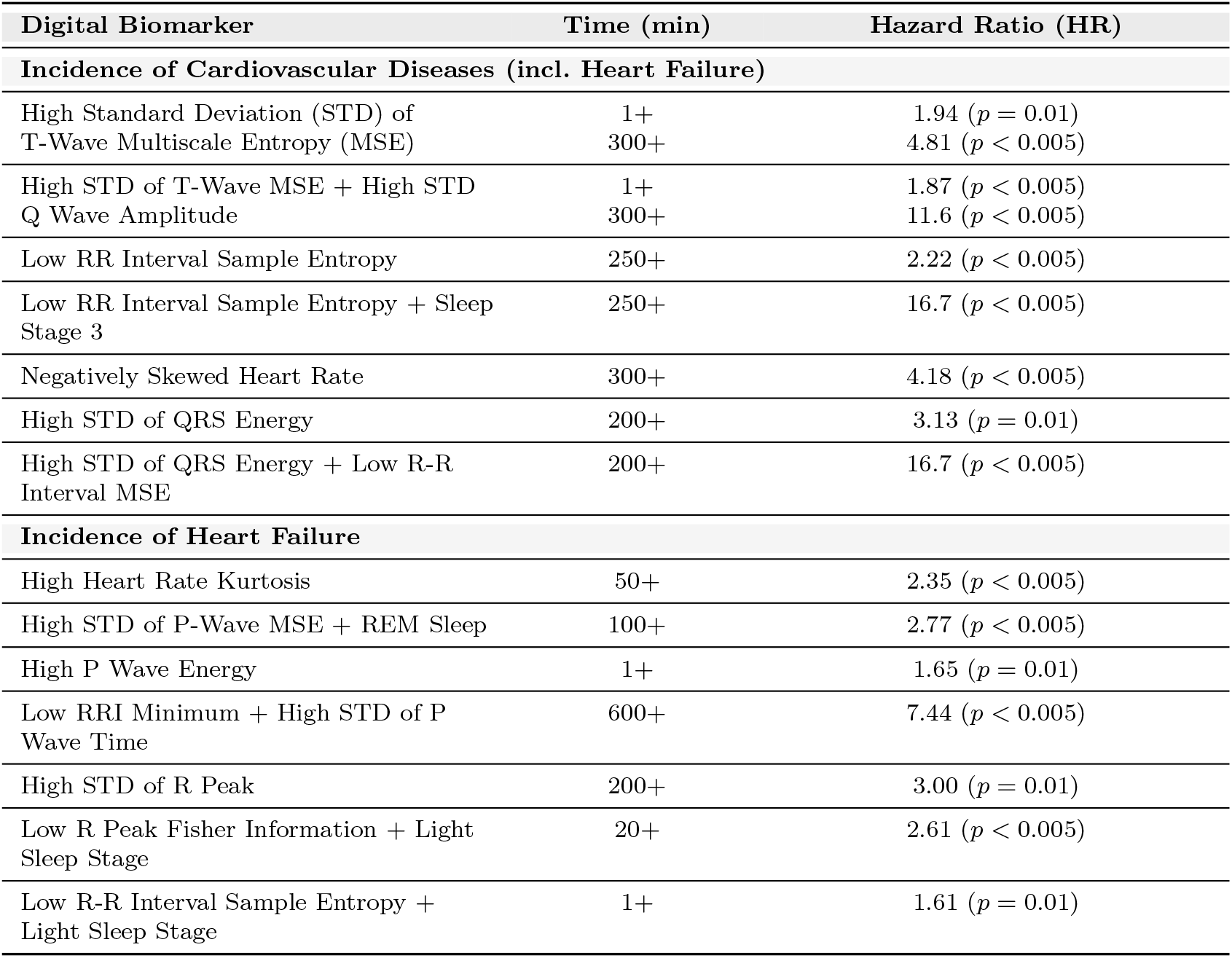
Overview of Digital Biomarkers found using the Digital Biomarker Discovery Pipeline.

| Digital Biomarker | Time (min) | Hazard Ratio (HR) |
| --- | --- | --- |
| **Incidence of Cardiovascular Diseases (incl. Heart Failure)** |  |  |
| High Standard Deviation (STD) of T-Wave Multiscale Entropy (MSE) | 1+<br>300+ | 1.94 ($p = 0.01$)<br>4.81 ($p < 0.005$) |
| High STD of T-Wave MSE + High STD Q Wave Amplitude | 1+<br>300+ | 1.87 ($p < 0.005$)<br>11.6 ($p < 0.005$) |
| Low RR Interval Sample Entropy | 250+ | 2.22 ($p < 0.005$) |
| Low RR Interval Sample Entropy + Sleep Stage 3 | 250+ | 16.7 ($p < 0.005$) |
| Negatively Skewed Heart Rate | 300+ | 4.18 ($p < 0.005$) |
| High STD of QRS Energy | 200+ | 3.13 ($p = 0.01$) |
| High STD of QRS Energy + Low R-R Interval MSE | 200+ | 16.7 ($p < 0.005$) |
| **Incidence of Heart Failure** |  |  |
| High Heart Rate Kurtosis | 50+ | 2.35 ($p < 0.005$) |
| High STD of P-Wave MSE + REM Sleep | 100+ | 2.77 ($p < 0.005$) |
| High P Wave Energy | 1+ | 1.65 ($p = 0.01$) |
| Low RRI Minimum + High STD of P Wave Time | 600+ | 7.44 ($p < 0.005$) |
| High STD of R Peak | 200+ | 3.00 ($p = 0.01$) |
| Low R Peak Fisher Information + Light Sleep Stage | 20+ | 2.61 ($p < 0.005$) |
| Low R-R Interval Sample Entropy + Light Sleep Stage | 1+ | 1.61 ($p = 0.01$) |

These findings not only showcase the predictive power of our approach but also provide meaningful insights into heart physiology, bridging the gap between advanced AI techniques and practical clinical use.

In summary, this paper has three contributions:

1. We introduce an *AI-based discovery pipeline* for interpretable digital biomarkers and apply it for the analysis of a large Cardiovascular Diseases (CVDs) dataset.
2. We show how our method successfully identifies both established biomarkers, such as *RR Interval Sample Entropy*, as well as novel biomarkers like *T-Wave Multi-scale Entropy*, which show strong associations with HF and other cardiovascular conditions.
3. We show how our findings *bridge the gap between advanced AI methodologies and clinical applicability* by providing physiologically meaningful insights for each biomarker, thereby promoting the integration of AI tools into clinical research and diagnosis.

## 2 Digital Biomarker Pipeline

Given an Electrocardiogram (ECG) signal, *E*(*t*), we extract a set of 314 interpretable features (e.g., Heart Rate Skew, P-wave amplitude, RR interval, etc.) that has proved to work well in ECG ML classification tasks [12, 13] from each 60-second segment of a single-lead ECG signal. This produces a two-dimensional array where one axis represents the interpretable ECG features and the other axis represents time.

Mathematically, the extracted features are represented as:

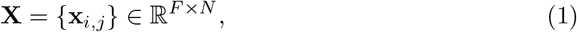

where, **x**_*i,j*_ is a scalar value corresponding to the *i*^*th*^ feature and *j*^*th*^ time segment. *F* is the number of extracted features, and *N* is the number of 60-second segments in the ECG recording.

For each segment *E*_*j*_(*t*) with *t* ∈ [*t*_*j*_, *t*_*j*_ + 60], the feature extraction function is:

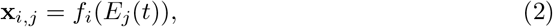

where *f*_*i*_(·) denotes the *i*^*th*^ feature extraction process applied to the *j*^*th*^ time segment.

This 2-D array **X** serves as the foundation for our analysis. Each ECG signal *E*(*t*) is associated with a patient group based on their clinical outcomes. Our primary goal is to identify anomalies within **X** that maximize the distinction between these patient groups. We hypothesize that these ECG biomarkers are present only during specific time intervals within the overall duration of the ECG recording. Consequently, employing a straightforward neural network to map each ECG signal *E*(*t*) to a patient group or to classify every 60-second segment is not feasible, as only a small subset of the data may contain the relevant information. This necessitates a more nuanced approach that can effectively isolate and analyze these informative segments to discover meaningful biomarkers. The overview of the proposed algorithm is shown in Fig. 1.

**Fig. 1:**
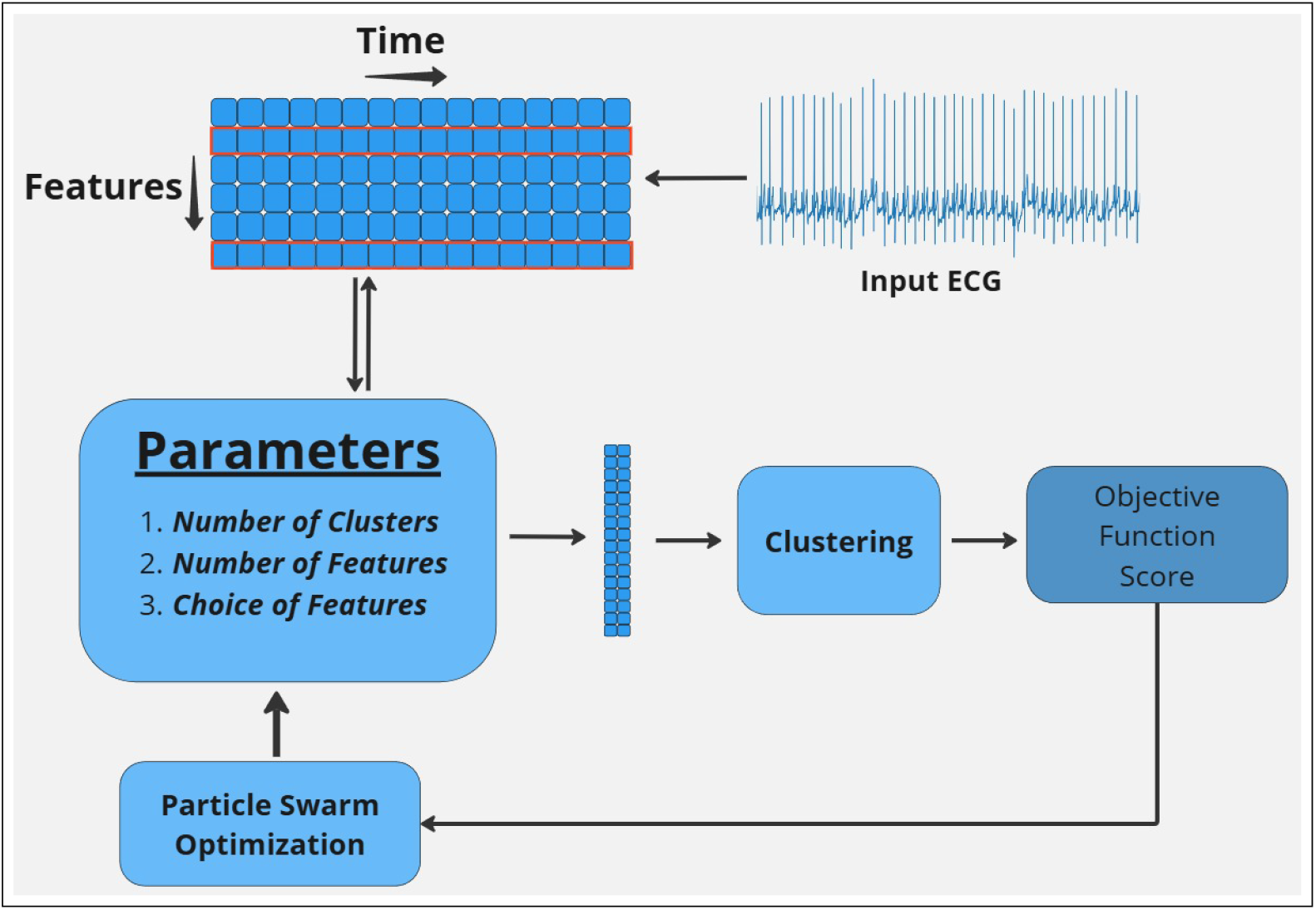
Overview of the proposed automated digital biomarker discovery pipeline for Cardiovascular Diseases (CVDs). The pipeline uses ECG signal preprocessing, feature extraction, and clustering of normalized features, an objective function to discover potential biomarkers and a Particle Swarm Optimization algorithm to automate the process.

### Analyzing Sub-sequences of ECG Data

We investigate sub-sequences of the ECG feature matrix **X** based on three hyperparameters:

- Cluster Size 𝒦 ∈ {30, 31, …, 120},
- Number of features *F*_1_ ∈ {1, 2, 3},
- Feature selection ℱ ⊆ {1, 2, …, *F*}.

Given the matrix **X** ∈ ℝ^*F* ×*N*^, we extract sub-sequences from features ℱ:

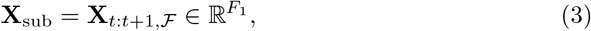

where ℱ is a subset of features, with length *F*_1_.

This process involves slicing the matrix **X** to obtain *N* segments across time and selecting specific features *F*_1_ from the total feature set ℱ. The choice of *F*_1_ and ℱ allows for targeted analysis of the data, enabling us to focus on specific feature combinations relevant to our study.

### Clustering of Latent Vectors

After obtaining sub-sequences **X**_sub_ for all patients, we apply an unsupervised learning algorithm, specifically K-Means clustering to group the representations. Each sub-sequence **X**_sub_ is assigned to a cluster label *c*_*j*_.

We then compute the cluster distributions for the risk group and the healthy group. For each cluster *k* ∈ 𝒦, where 𝒦 is the set of clusters, we define the proportions of risk and healthy patients assigned to the cluster as:

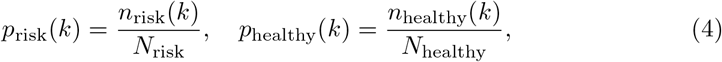

where *n*_risk_(*k*) and *n*_healthy_(*k*) are the number of risk and healthy patients assigned to cluster *k. N*_risk_ and *N*_healthy_ are the total number of risk and healthy patients, respectively.

### Quantifying Cluster Differences

To quantify the difference between the cluster distributions in the HF and healthy groups, we define the objective function Δ_max_ as:

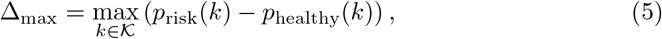

which identifies the cluster with the largest difference between the two groups. We also introduce an adjusted objective function Δ_adj_, which penalizes clusters common in the healthy group:

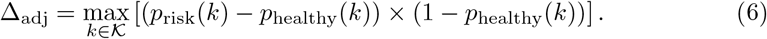

This adjustment emphasizes clusters that are prevalent in the HF group but less frequent in the healthy group.

### Optimization of Parameters

In our search for digital biomarkers, we hypothesize that multiple potential biomarkers exist in the high-dimensional feature space, each with different levels of efficacy in distinguishing high-risk patients. However, an exhaustive search evaluating all parameter combinations would be computationally infeasible. Based on our calculations, performing a complete search would require approximately 883 years on a 16GB RAM computer, as each configuration takes roughly one minute to compute (though this is dependent on cluster size). This estimate derives from the combinatorial space: with ^314^*C*_3_ feature combinations, 90 possible cluster sizes, and 30 time length settings, resulting in:

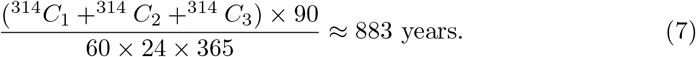

To address this complexity and maximize the objective functions for different combinations of hyperparameters *T*_1_, *C*_1_, *F*_1_, and ℱ, we model this task as an state-space optimization problem. We use Particle Swarm Optimisation (PSO) for this task due to its alignment with the objectives of our biomarker discovery pipeline: Our approach requires identifying multiple biomarkers potentially spread throughout the search space, rather than isolating a single global optimum. While PSO is known to occasionally converge prematurely to local optima in high-dimensional spaces [14], we leverage this characteristic by running multiple, rapid experiments with varied initial conditions to capture a broad range of biomarkers across local minima. PSO’s efficient balance of exploration and exploitation enables it to navigate complex feature spaces effectively, providing swift convergence without excessive computational demands.

By maximizing these metrics, we aim to uncover clusters associated with a high risk of target disease, potentially revealing novel digital biomarkers that differentiate patients at risk from healthy controls. This analysis helps isolate critical time intervals and features relevant for early detection and intervention in CVDs.

## 3 Identification of Digital Biomarkers in CVD

We apply the digital biomarker discovery pipeline method described in Section 2 to find digital biomarkers from the Sleep Heart Health Study (SHHS) dataset. Table 1 provides an overview of the identified digital biomarkers for identifying incidences of CVDs and HF, respectively. Note that HF is included in the CVDs class, but since HF is such an important type of cardiovascular incidence, we identify the digital biomarkers for HF separately. The more robust digital biomarkers are described in detail in the main part of this paper, while the explanation of additional biomarkers is found in Appendix A.

### 3.1 Dataset

For all analyses conducted in this study, we utilized the SHHS dataset [15], a multicenter cohort comprising polysomnographic data from 6441 participants along with their follow-up clinical outcomes. This is an existing database and access for this dataset can obtained via the online portal: www.sleepdata.org. For each participant in the dataset, we extracted the ECG signals recorded throughout their entire night’s sleep, as well as clinical variables such as age, gender, race, and CVD outcomes. The ECG signals were preprocessed by applying a bandpass (0.5 - 40Hz) filter and subsequently normalized. To facilitate the biomarker discovery process, we randomly divided the dataset into two equal halves, serving as the discovery and validation cohorts, respectively.

### 3.2 Incidence of CVDs

The analysis of CVDs encompasses the following conditions: HF, Stroke, and Myocardial Infarction (MI). We define the incidence of CVDs as the occurrence of a new diagnosis of CVDs during the follow-up period among patients who had no diagnosed CVDs at baseline. The control group is not diagnosed with CVDs at baseline nor follow-up. For this configuration, we have identified the following digital biomarkers.

#### 3.2.1 T-Wave Multiscale Entropy STD

##### Definition

T-wave multiscale entropy measures the complexity of ventricular repolarization dynamics over various time scales, capturing both short-term and long-term patterns.

##### Clinical Significance

Diseases like ischemia, cardiomyopathy, or electrolyte imbalances can alter repolarization complexity. Hyperacute T-waves are early signs of acute MI and represent a significant deviation from normal T-wave morphology [16].

##### Findings

The proposed method finds that a high STD (*>*0.25) of T-Wave Multi-scale Entropy (as seen in Fig. 2a) is associated with CVD even if a single instance of this is present. Hazard Ratio (HR) = 1.94 (95% CI: 1.22–3.08, *p* = 0.01), and if this anomaly continues (300+ minutes), its risk increases to Hazard Ratio (HR) = 4.81 (95% CI: 1.94–11.95, *p <* 0.005). Interestingly, we also find that if this occurs simultaneously with a high STD (*>* 0.06) of **Q Wave Amplitude** (Fig. 2b), we see a risk of **Hazard Ratio (HR)** = 1.87 **(95% CI: 1.22–2.89**, *p <* 0.005) for a single instance and **Hazard Ratio (HR)** = 11.60 **(95% CI: 2.77–48.55**, *p <* 0.005) when it persists for 300+ minutes.

**Fig. 2:**
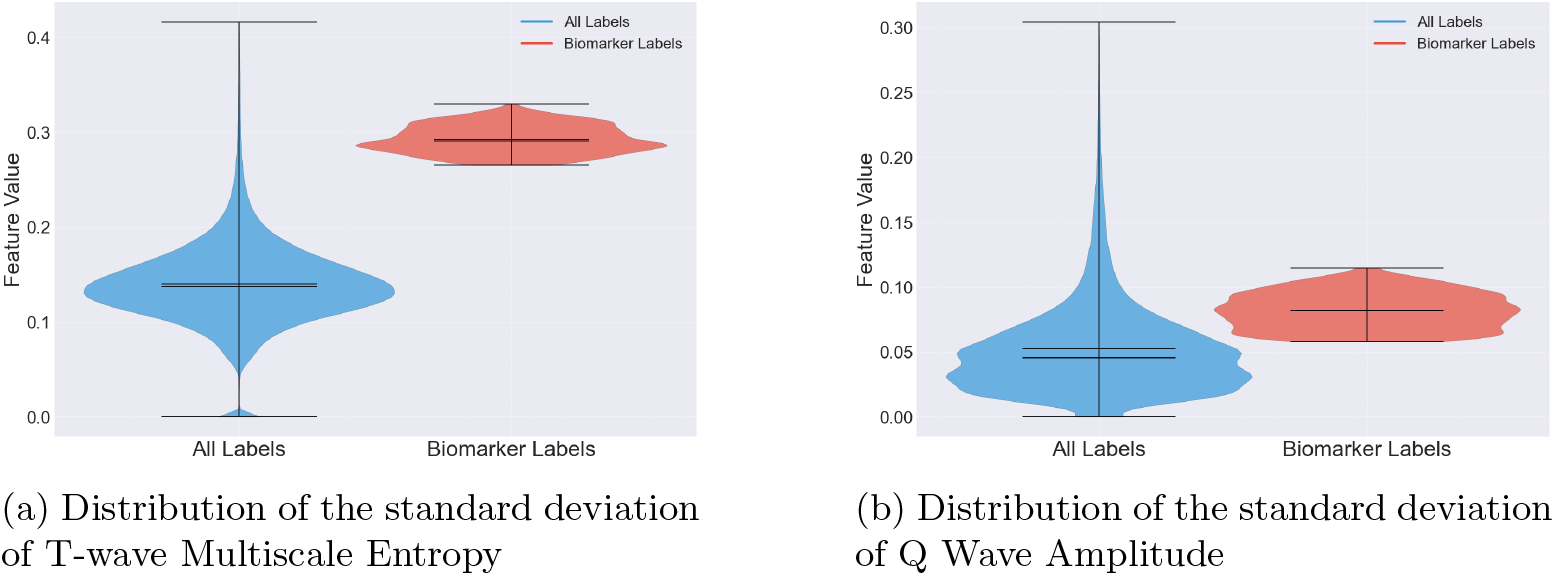
Comparison of biomarker distribution between patients at risk of CVD and healthy individuals.

##### Existing Literature

T-Wave Alternans (TWA) can be seen as a similar feature of T-Wave Multiscale Entropy STD and has been shown to be a predictor of Sudden Cardiac Death [17–19] and post-operative cardiac mortality [20]. Studies have assessed the risk of multiscale entropy of Heart Rate Variability (HRV) in relation to CVD [21]. However, no studies have demonstrated an association between T-wave multiscale entropy during high STD of Q-wave amplitude and the incidence of HF.

##### Physiological Justification

Diseases such as ischemia and cardiomyopathy disrupt the uniformity of ventricular repolarization, introducing heterogeneity in myocardial electrical activity [22]. This disruption increases the complexity and variability of the T-wave morphology over multiple time scales. Multiscale entropy quantifies this complexity; a high STD of T-wave multiscale entropy reflects significant fluctuations in repolarization dynamics, which might be associated with cardiovascular dysfunction. Therefore, elevated T-wave multiscale entropy STD can serve as a physiological marker for underlying CVD.

#### 3.2.2 RR Interval Sample Entropy

##### Definition

RR Interval Sample Entropy measures the unpredictability and complexity of heart rate dynamics, offering insights beyond traditional linear HRV metrics. A lower sample entropy value suggests increased regularity and predictability in heart rate, indicating a more homogeneous and less complex system.

##### Clinical Significance

Low RR Interval Sample Entropy is associated with reduced variability and increased regularity in heart rate patterns. This reduction in complexity is often linked to aging, CVD, and conditions such as HF. A more predictable heart rate may reflect an underlying pathological state, as healthy heart dynamics tend to exhibit more variability and adaptability.

##### Findings

In our study, we found that a low RR Interval Sample Entropy (*<* 1.5) during sleep was associated with a significantly higher risk of CVD. Specifically, individuals with this metric persisting for more than 250 minutes in a night had a **Hazard Ratio (HR) of 2.22** (95% CI: 1.29–3.83, *p <* 0.005). These findings suggest that lower sample entropy could serve as an indicator of increased risk for age-related cardiovascular issues. Moreover, the risk increased significantly when low RR Interval Sample Entropy was observed during **Sleep Stage 3**, with a **Hazard Ratio (HR) of 16.70** (95% CI: 3.97–70.26, *p <* 0.005), suggesting that this metric may be a more robust biomarker during this specific sleep phase.

##### Previous Literature

Our observation is consistent with previous studies that have shown older adults exhibit higher RR Interval Sample Entropy values compared to younger individuals, indicating a greater degree of irregularity in their heart rhythms. The findings imply that lower sample entropy values could indicate an increased risk for age-related cardiovascular issues [11, 23]. There have also been findings correlating deep sleep with low RRI entropy in Post-MI patients [24], but no studies have explored this as a biomarker for CVDs.

##### Physiological Justification

During Sleep Stage 3, known as deep sleep, the body undergoes increased parasympathetic activity, leading to greater variability and complexity in heart rate dynamics under normal conditions [24]. A low RR Interval Sample Entropy during this stage indicates reduced heart rate complexity and adaptability to physiological fluctuations. This reduction suggests autonomic dysfunction, a common feature in CVD, where impaired parasympathetic activity and unopposed sympathetic dominance decrease HRV. Therefore, low RR Interval Sample Entropy during Sleep Stage 3 serves as a robust biomarker for underlying cardiovascular dysfunction, reflecting the heart’s diminished capacity to respond to physiological needs during a critical restorative phase.

### 3.3 Incidence of Heart Failure (HF)

We define a Risk and Healthy group similar to the above experiment but only accounting for HF cases.

#### 3.3.1 High Heart Rate Kurtosis

##### Definition

Kurtosis measures the degree to which a distribution is peaked or flat compared to a normal distribution. A higher kurtosis indicates a very narrow distribution and also suggests a higher occurrence of extreme HRV.

##### Clinical Significance

HRV reflects the balance between the sympathetic and parasympathetic nervous systems. A high heart rate kurtosis may indicate a dominance of one autonomic branch leading to extreme HRV. This could also be associated with stress, anxiety, or autonomic dysfunction.

##### Findings

It is found that a very high heart rate kurtosis (*>*10) sustained over a prolonged period (*>*50 minutes), is indicative of HF with a **Hazard Ratio (HR)** = 2.35 **(95% CI: 1.38–4.01**, *p <* 0.005) or a 1% increase in risk for every minute of high heart rate kurtosis *p <* 0.005. This is contrary to the biomarker for CVD mentioned in Section 3.2.2. This is because as kurtosis increases, outliers in the distribution increase too, thereby increasing RR interval sample entropy. While we find that a low (*<* 1.5) RR Interval Sample Entropy is a biomarker for CVD, we do not find this association for HF. Instead, we find an association if the RR Interval Sample Entropy is low but greater than 0 (0*<*x*<*1.5) with Hazard Ratio (HR) = 1.51 (95% CI: 1.03–2.21, *p* = 0.03). However, we also find a strong association for HF if the heart rate kurtosis is high (*>* 10) (as seen in Fig. 3a) and the RRI Sample Entropy is low (*<*1.5) (Fig. 3b) in the same one-minute recording. We find an increase of risk by 2% for every minute a patient spends with this heart rhythm (*<*0.005).

**Fig. 3:**
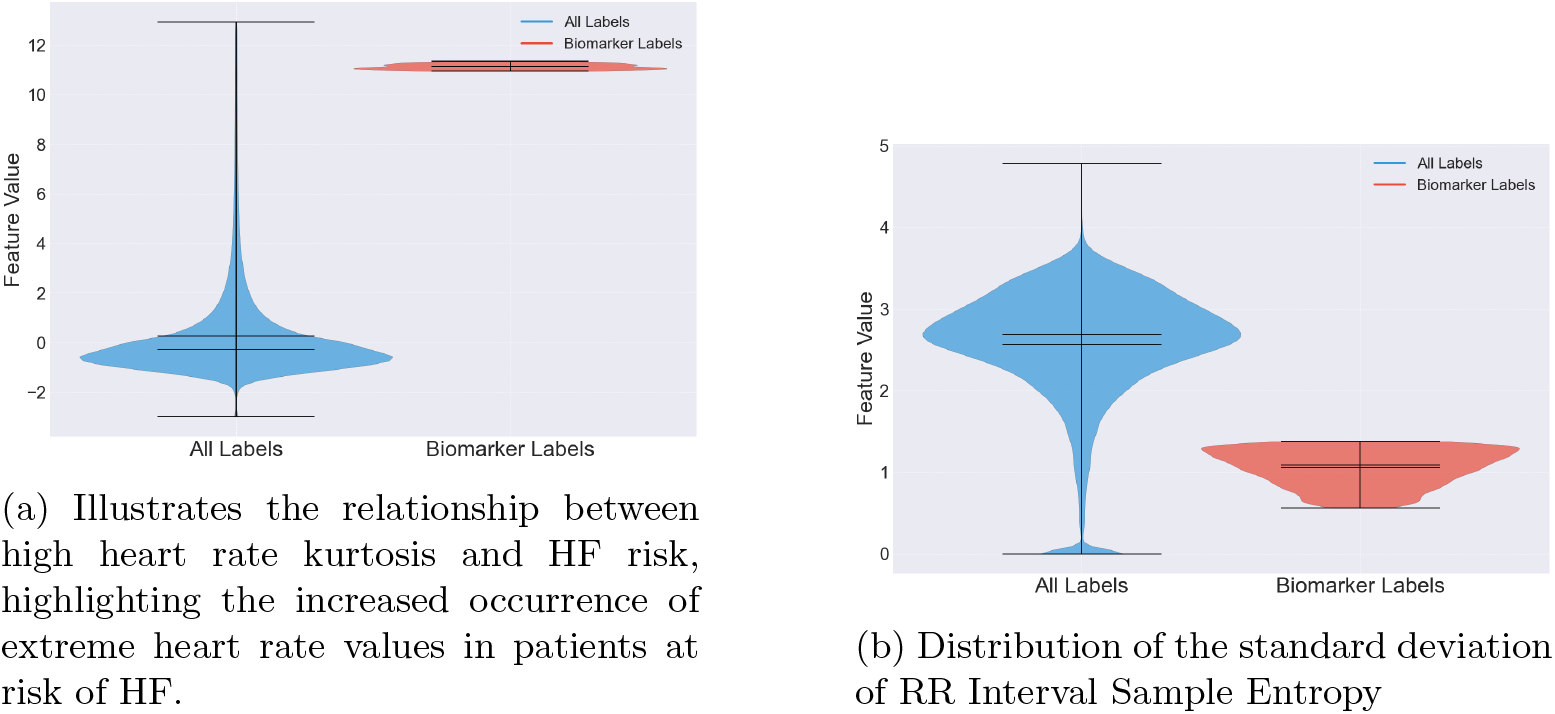
Distribution of key biomarkers associated with Heart Failure (HF) across the discovery cohort.

##### Existing Literature

Previous studies [25, 26] discuss how a consistently high resting heart rate is linked to cardiovascular risk and also its association with white matter abnormalities. But, no studies map high heart rate kurtosis to HF. It should also be noted that the assocciation we found is independent of the mean heart rate, suggesting that outliers, even at low resting heart rates, can be dangerous.

##### Physiological Justification

HF often involves structural changes in the myocardium, such as fibrosis and ventricular dilation. These changes contribute to electrical heterogeneity and instability, predisposing patients to arrhythmias. The combination of kurtosis to low sample entropy reflects a compromised autonomic system that cannot adequately buffer sudden cardiac stressors, increasing the risk of adverse outcomes.

#### 3.2.2 P-Wave Multiscale Entropy (STD)

##### Definition

The STD of P-Wave Multiscale Entropy quantifies the variability of entropy values across different scales or over time, reflecting the consistency or variability in atrial depolarization complexity.

##### Clinical Significance

Abnormalities in P-Wave MSE STD may reflect dysregulation of autonomic control over the heart, which can be important in conditions like sleep apnea or HF.

##### Findings

We find that a high STD of P-Wave Multiscale Entropy (*>*0.3) alone is not a reliable marker for HF with a **Hazard Ratio (HR) of 1.37** (95% CI: 0.97–1.92, *p* = 0.07). However, we see that this condition during **REM sleep** increases the risk of HF adding a 1% increase in risk for every 1 min spent with a high STD (*p <* 0.005). We also find that a sustained state for 100+ minutes increases the risk with a **Hazard Ratio (HR) of 2.77** (95% CI: 1.56–4.92, *p <* 0.005).

##### Existing Literature

While there has been studies about multiscale entropy analysis on HRV [27] and also during sleep stages [28], there has been insufficient works describing the behaviour of P-wave especially during REM sleep.

##### Physiological Justification

An elevated P-wave multiscale entropy standard deviation (MSE STD) during REM sleep is an anomaly that indicates increased variability and complexity in atrial depolarization dynamics, not typically seen in healthy individuals. This abnormality reflects disrupted atrial electrical activity and conduction heterogeneity, which are hallmark features of HF. In HF, structural changes like atrial dilation and fibrosis lead to irregular atrial conduction. During REM sleep, heightened autonomic fluctuations exacerbate these irregularities, resulting in significant deviations in P-wave complexity. Therefore, a high P-wave MSE STD during REM sleep might serve as a physiological indicator of HF.

## 4 Discussion

### 4.1 Main Findings

We have presented the Digital Biomarker Discovery Pipeline, which is a novel AI-driven approach for discovering digital biomarkers from ECG data to predict CVDs, particularly HF. By leveraging interpretable feature extraction and clustering techniques, we identified explainable digital biomarkers that differentiate between patient groups. The inclusion of PSO enabled efficient hyperparameter tuning and an automated approach to the pipeline.

A study of ECG data using this discovery pipeline identified several strong digital biomarkers in CVD, including ‘RR Interval Sample Entropy’ and the ‘T-wave Correlation Coefficient Median’ and their associations with sleep stages, thereby demonstrating strong associations with the incidence of HF and other cardiovascular conditions. These biomarkers provide clinically relevant insights that could contribute to early detection and intervention.

The proposed pipeline not only uncovers significant digital biomarkers but does so in a way that is both interpretable and clinically actionable. This work paves the way for integrating AI-based methods into clinical practice, enhancing preventive healthcare by allowing for more precise and timely identification of patients at risk of cardiovascular events. Future work will focus on validating these biomarkers across larger, more diverse cohorts and extending the approach to include additional cardiovascular conditions.

### 4.2 Limitations

While Particle Swarm Optimisation (PSO) substantially reduces the search time for hyperparameters, it is not without limitations. One primary drawback is that, by using PSO rather than an exhaustive search, there is an inherent risk of missing potential biomarkers, as only a portion of the full parameter space is explored. This means that there is a possibility that some biomarkers with valuable predictive power may remain undiscovered if they lie outside the regions explored by the algorithm.

Another limitation is PSO’s lack of memory across runs, which increases the likelihood of re-discovering biomarkers identified in previous runs. Without an overarching memory mechanism to retain previously found solutions, the algorithm may revisit similar regions of the search space, potentially reducing the efficiency of each new run in identifying novel biomarkers. These issues highlight areas for future optimization, where integrating a memory component or adaptive exploration-exploitation strategies could further improve biomarker discovery.

## Data Availability

The SHHS dataset used in this study has
been archived by the National Sleep Research Resource [15] with appropriate deidentification.
Permissions and access for these datasets were obtained via the online
portal: www.sleepdata.org.

## List of Acronyms

ECG: Electrocardiogram
HF: Heart Failure
CVD: Cardiovascular Disease
AI: Artificial Intelligence
PSO: Particle Swarm Optimisation
HR: Hazard Ratio
MSE: Multiscale Entropy
STD: Standard Deviation
SHHS: Sleep Heart Health Study
HRV: Heart Rate Variability

## Acknowledgments

This research has been funded by the Innovation Fund Denmark as part of the CATCH project (Project No. #1061-00046B) and the Copenhagen Center for Health Technology (CACHET). The funders played no role in the analysis or the writing of this manuscript.

## Declarations

### Funding

This research has been funded by the Innovation Fund Denmark as part of the CATCH project (Project No. #1061-00046B) and the Copenhagen Center for Health Technology.

### Conflict of interest

The author declares no competing interests.

### Availability of data and materials

The SHHS dataset used in this study has been archived by the National Sleep Research Resource [15] with appropriate de-identification. Permissions and access for these datasets were obtained via the online portal: www.sleepdata.org.

### Code availability

The underlying code for this study is not publicly available but may be made available to qualified researchers on reasonable request to a corresponding author.

### Authors’ contributions

All authors were part of conceptualizing and designing the work. GM collected the data, designed and implemented the methods, and carried out the data analysis. All authors were part of the interpretation of the results. GM wrote the first draft of the manuscript and all authors contributed to further revisions. All authors have approved the final version of the manuscript. JEB supervised the project. HD, JEB, and SP acquired funding for the project.

## A Appendix

While we found a multitude of different digital biomarkers both independently or a combination of them associated with the target outcomes, we highlight and explain a few more notable biomarkers in this appendix.

### A.1 Incidence of Heart Failure (HF)

#### A.1.1 High P Wave Energy

P wave energy refers to a quantitative measure of the electrical activity associated with atrial depolarization.

Figure. 4 shows the distribution of P Wave Energy across the full training set, and our method shows a small area that stands out between the two groups. Using this information, we check if **High P Wave Energy (***>* **6)** has predictive capabilities. On applying this finding to the testing cohort, we find this marker to have a **Hazard Ratio (HR)** = 1.65 **(95% CI: 1.16–2.34**, *p* = 0.01).

**Fig. 4:**
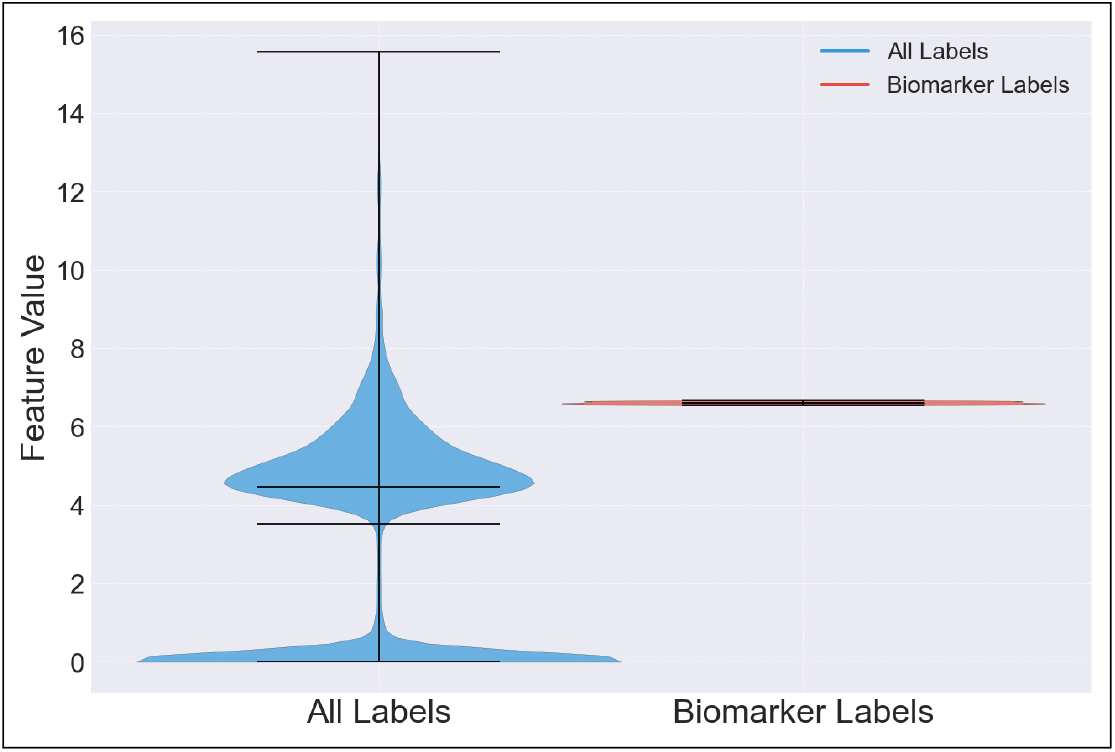
Violin plot showing the distribution of P Wave Energy across patients with risk and healthy controls. The plot highlights an increase in P Wave Energy in patients at risk of Heart Failure (HF), suggesting that this biomarker may serve as an indicator of atrial depolarization abnormalities associated with HF.

#### A.1.2 RR Interval-P Wave Time

We find that a Low RRI Minimum (*<* 0.5) (defined as the lowest RR Interval in a 60-second segment) and a High Standard Deviation of P Wave Time (*>* 0.0001) when it occurs simultaneously and persistently (*>* 600*minutes*), has a **Hazard Ratio (HR)** = 7.44 **(95% CI: 2.29–24.16**, *p <* 0.005). Although, a Low RRI Minimum occurring independently, does not seem to be a risk factor Hazard Ratio (HR) = 1.84 (95% CI: 0.68–5.00, *p* = 0.23)

#### A.1.3 R Peak Metrics

The proposed method shows that a **high standard deviation (***>* 0.2**) of R Peak** sustained over a long period (more than 200 minutes) has a Hazard Ratio (HR) = 3.00 (95% CI: 1.32–6.85, *p* = 0.01). It is also observed that a **Low R Peak Fisher Information during Light Stage Sleep** for more than 20 minutes has a Hazard Ratio (HR) = 2.61 (95% CI: 1.42–4.77, *p <* 0.005) or a 2% increase in risk for every minute in this state. This risk is only observed during Light Stage sleep and R Peak Fisher Information is not independently associated with HF Hazard Ratio (HR) = 0.99, *p* = 0.25). We also find that a **low RR Interval Sample Entropy** (*<*1.5) during Light Sleep is a risk if it exists for more than a minute Hazard Ratio (HR) = 1.61 (95% CI: 1.11–2.34, *p* = 0.01).

### A.2 Incidence of Cardiovascular Diseases (CVDs)

#### A.2.1 Heart Rate Metrics

It is observed that a **negatively skewed heart rate** (*<*-2) persisting for more than 300 minutes increases cardiovascular risk Hazard Ratio (HR) = 4.18 (95% CI: 1.67–10.49, *p <* 0.005).

### A.3 QRS Energy STD - RR Interval MSE

We find that a high Standard Deviation (STD) of QRS Energy (*>* 2) occurring simultaneously with a low Multiscale Entropy (MSE) of R-R Interval (*<* 1.2) is strongly associated with the incidence of CVD with an increase in the risk of 1% for every minute. If this anomaly is sustained for more than 200 minutes, we find a risk of Hazard Ratio (HR) = 16.70 (95% CI: 3.97–70.26, *p <* 0.005). We also find that a high Standard Deviation (STD) of QRS Energy is independently associated with CVD with a Hazard Ratio (HR) = 3.13 (95% CI: 1.37–7.15, *p* = 0.01) and also a low RRI MSE with a Hazard Ratio (HR) = 2.14 (95% CI: 1.22–3.75, *p* = 0.01) if they are sustained for more than 200 minutes.

## Notes

### Competing Interest Statement

The authors have declared no competing interest.

